# Uncovering Neural Signatures of Convulsive Therapy in Depression Using Massive EEG Time-Series Feature Extraction

**DOI:** 10.64898/2026.02.05.26345687

**Authors:** Aron T. Hill, Kate Godfrey, Jarrad A.G. Lum, Reza Zomorrodi, Daniel M. Blumberger, Paul Fitzgerald, Zafiris J. Daskalakis, Neil W. Bailey

## Abstract

**BACKGROUND:** Convulsive therapies, including electroconvulsive therapy (ECT) and magnetic seizure therapy (MST), are highly effective for treatment-resistant depression, however, their neural mechanisms remain incompletely understood. We tested whether a data-driven framework applying a massive time-series feature library to electroencephalography (EEG) could reveal novel insights into changes in functional brain dynamics following convulsive therapy and provide preliminary markers of response.

**METHODS:** Resting-state EEG was analysed before and after a course of ECT or MST in 42 patients. Data were pooled and reduced to three principal components (PCs) capturing 78.1% of total variance, then >7,000 time-series features per PC were extracted from each patient using the highly comparative time-series analysis (*hctsa*) framework. Linear support vector machines (SVMs) classified pre-versus post-treatment EEG for each PC. A separate linear SVM, was also trained on 18 representative baseline features to predict clinical response.

**RESULTS:** *hctsa-*based classifiers distinguished pre-from post-treatment EEG for each PC (accuracy 73.8-76.2%, p_FDR_<0.01). Between 986-1,414 features significantly differentiated post-stimulation from baseline time-series across the three PCs (p_FDR_<0.05). The most discriminative features indexed linear and non-linear autocorrelation, correlation, multiscale entropy, and spectral properties. The baseline SVM combining 18 features showed modest but statistically reliable prediction of treatment response (balanced accuracy=0.69, area under the curve [AUC]=0.61, p=0.014). Single-feature ROC analyses further identified several top features with AUCs=∼0.7.

**CONCLUSIONS:** Data-driven analysis using a diverse time-series feature library can uncover novel EEG signatures of brain changes following convulsive therapy. This holds potential for delineating treatment-related mechanisms and developing predictive biomarkers to support precision psychiatry.

## Introduction

Convulsive therapies, including electroconvulsive therapy (ECT) and magnetic seizure therapy (MST) are efficacious device-based interventions for severe refractory psychiatric illness, most notably treatment resistant depression [1]. Both interventions intentionally induce therapeutic seizures but differ in how stimulation is administered. ECT applies an electrical current across the scalp to produce a generalised seizure, whereas MST uses high-frequency, rapidly alternating magnetic fields that target cortical regions with more focal stimulation to produce a generalised seizure [2, 3]. Convulsive therapies have demonstrated robust antidepressant effects, with response rates between approximately 40-70% [4-7]. However, despite their clinical efficacy, the mechanisms by which therapeutic seizures alleviate depression remain poorly understood.

Electroencephalography (EEG) has long been used to characterise functional brain changes following convulsive therapies for major depressive disorder (MDD) [8, 9]. The most consistent post-seizure finding is a shift towards slower oscillatory activity, typically involving increased theta and delta power following treatment [8, 10, 11]. This slowing has been reported both qualitatively via visual inspection of EEG recordings [12, 13], and quantified using power spectral analyses [14-17]. Additional work has begun to extend these observations to more complex markers of brain dynamics, including treatment-related changes in functional connectivity [14, 17-19], large-scale network activity (via EEG microstates) [20], and broadband aperiodic activity [11, 21]. However, these measures capture only a narrow portion of the complex, multidimensional EEG signal, leaving other potentially informative aspects of neural dynamics uncharacterised, including aspects that may index treatment-induced changes in neural activity following convulsive therapy.

To better understand how convulsive therapies modulate neural activity in MDD, we sought to apply a highly comparative time-series analysis (*hctsa*) framework [22]. *hctsa* extracts over 7,000 univariate time-series features derived from an extensive interdisciplinary library of algorithms encompassing physics, biology, neuroscience, biomedicine, economics, and engineering [22]. These features capture fundamental statistical properties of the distributions of time-series, their linear correlations across time points, stationarity measures, and entropy-based metrics, among numerous others. A key benefit of this approach is that it enables systematic, large-scale comparison of many time-series features that would be impractical to analyse individually using a hypothesis-driven approach. This data-driven strategy may therefore reveal novel features and previously unrecognised mechanisms that are difficult to detect using traditional methods.

The *hctsa* library has been applied in diverse scientific and clinical contexts, such as identifying relevant time-series features associated with psychiatric disorders [23], distinguishing healthy EEG patterns from epileptic seizures [24], and predicting responders to transcranial magnetic stimulation (TMS) treatment for depression [25]. Here, we sought to capitalise on the extensive feature library provided by *hctsa*, with the aim to identify key changes in brain dynamics following convulsive therapy for depression (combining EEG data from two separate ECT and MST studies [26, 27]). Discovering features that reliably distinguish post-treatment from pre-treatment activity could offer mechanistic insights into how these interventions modulate neural dynamics.

## Methods and Materials

For brevity, detailed information is provided in the Supplemental Information, with key information summarised here.

### Datasets

Data were drawn from two previously conducted open-label studies in MDD; an MST trial [26] (ClinicalTrials.gov identifier: NCT01596608; CONSORT Diagram in Supplementary Figure S1) and an ECT study [27]; both conducted at the Centre for Addiction and Mental Health (CAMH). All participants provided written informed consent, and protocols were approved by the CAMH Research Ethics Board in accordance with the Declaration of Helsinki. Findings from these studies have been reported previously [e.g., 14, 17, 20, 21, 28]. Here, we apply a unique data-driven analysis approach to these datasets.

### Participants

Data from a total of 42 participants (21 ECT, 21 MST) were included in analyses after removal of three participants with insufficient clean baseline or post-treatment EEG data. All participants had a diagnosis of MDD based on DSM-IV-TR criteria. Participant demographics are provided in Table 1.

**Table 1.**
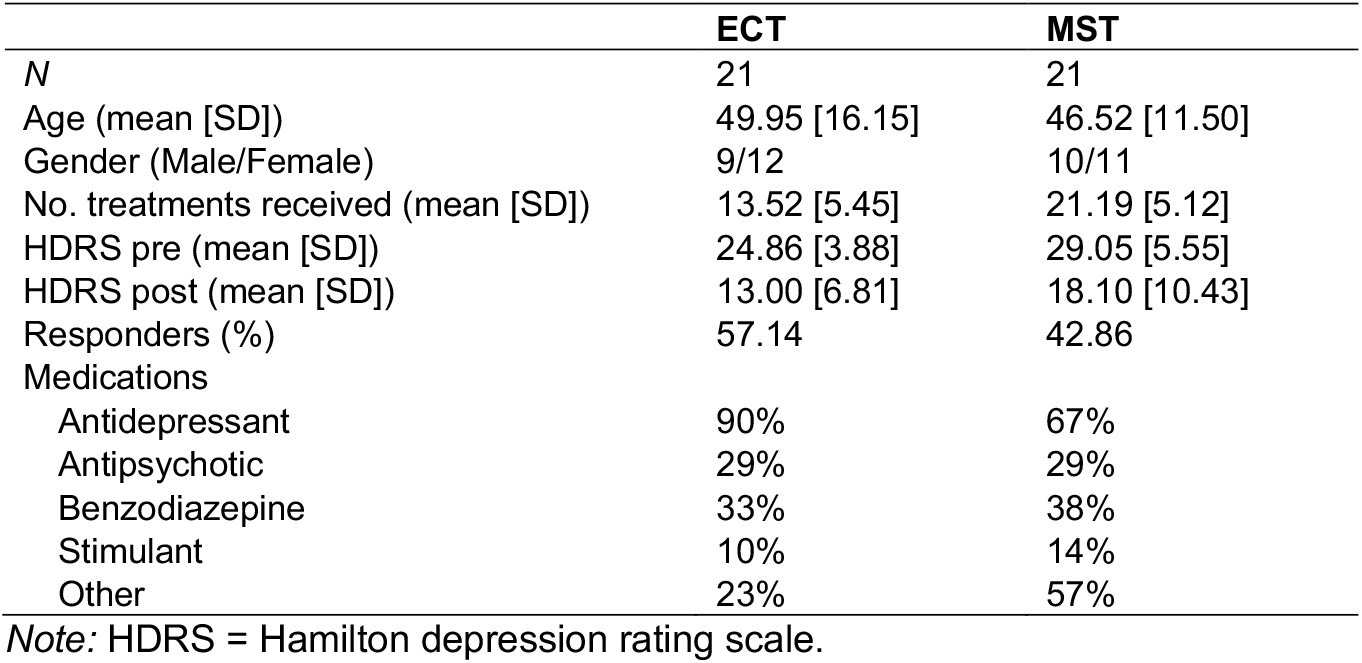
Demographics and clinical characteristics of participant included in analysis.

### Convulsive Therapy

ECT was administered 2-3 times per week using a brief-pulse MECTA device. Stimulation was predominantly right unilateral ultra-brief (0.3 ms) with bitemporal placement as required. Electrode positioning followed APA guidelines [27, 29]. MST was delivered 2-3 times per week (MagPro MST stimulator; bilateral Twin Coil-XS over F3/F4, targeting dorsomedial prefrontal cortex), until remission (HDRS-24 score ≤ 10 and ≥60% symptom reduction on two consecutive ratings) or a maximum of 24 sessions [26]. Sessions were conducted under general anaesthesia with neuromuscular blockade (detailed protocol in Supplement; and [26, 27]).

### EEG Data Acquisition and Pre-Processing

Ten minutes of eyes-closed resting EEG were recorded using a 64-channel Neuroscan Quik-cap and SynAmps2 amplifier (Compumedics, USA) with 0.05-1,000 Hz online bandpass filtering and vertex reference. Baseline EEG was acquired within the week before treatment, and post-treatment EEG was obtained within 2 days for ECT and approximately four days for MST. Data were pre-processed in MATLAB using the automated RELAX v2.0.0 pipeline (full details in Supplement) [30-32]. Clean data were segmented into 30 s epochs, consistent with previous work [33, 34] and the first artefact-free epoch per participant was analysed, pooling ECT and MST to maximise power.

Data were down-sampled to 160 Hz and z-score normalised across electrodes and timepoints within each epoch to minimise amplitude differences across participants while preserving relationships between electrodes [33]. As EEG data are multivariate (i.e., 60 time-series, one per electrode) and *hctsa* requires a single univariate time-series, consistent with prior studies, we applied principal component analysis (PCA), to reduce EEG data to the top three PCs that captured significant variance in the EEG data [25, 33]. Prior to PCA, the time-series data were concatenated across participants in the temporal dimension to ensure all PCA weights matched across participants [33, 35], while the z-score transform noted earlier prevented potential class imbalances in EEG amplitudes from disproportionately influencing PCA components towards individuals with larger signals. The top 3 PCs (explaining 78.15% variance) were selected for further analysis.

### hctsa Feature Extraction

*hctsa* (version 1.09) [22, 24] was implemented in MATLAB (R2021a) to perform a massive feature extraction of the EEG-derived time-series for each PC. Any non-real values or errors returned from the feature extraction were excluded from further analysis. A feature was excluded if it yielded non-realistic or erroneous outputs for all participants. This resulted in between 7,093-7,119 features for each of the three PCs. All participants provided real values for all features. A comprehensive list of feature counts is available in the Supplement (Table S1). A robust scaled mixed sigmoid transformation was subsequently applied to normalise the features, enabling meaningful comparisons across outputs with varying ranges and distributions [24].

After normalisation, we used a linear support vector machine (SVM) classifier (MATLAB *fitcsvm*, linear kernel, default regularisation, no hyperparameter tuning) to discriminate baseline from post-treatment conditions for each PC based on the *hctsa* feature set, implementing a leave-pair-out cross-validation approach for classification performance testing, i.e., holding out on each fold one matched-pair consisting of a participant’s baseline and post-treatment data, and training on the remainder of the data. As such, fold accuracy could only be 0, 50, or 100 percent. We report the mean across all fold accuracies. Because each fold’s test set was balanced, this quantity is numerically identical to balanced accuracy. While computationally more intensive than other methods such as *k*-fold cross-validation, this method can aid in maximising the use of limited data by iteratively training on all but one participant’s baseline and post-treatment pair, and then testing on the excluded pair, thereby providing an unbiased estimate of model performance [36, 37]. To assess statistical significance and ensure results were not positively biased due to model overfitting, a null distribution of accuracies was generated by randomly permuting the pre- or post-treatment labels associated with each EEG file and re-training the SVM on each permuted dataset (total of 1000 permutations). A graphical overview of the overall analysis approach using *hctsa* is provided in Figure 1.

**Figure 1.**
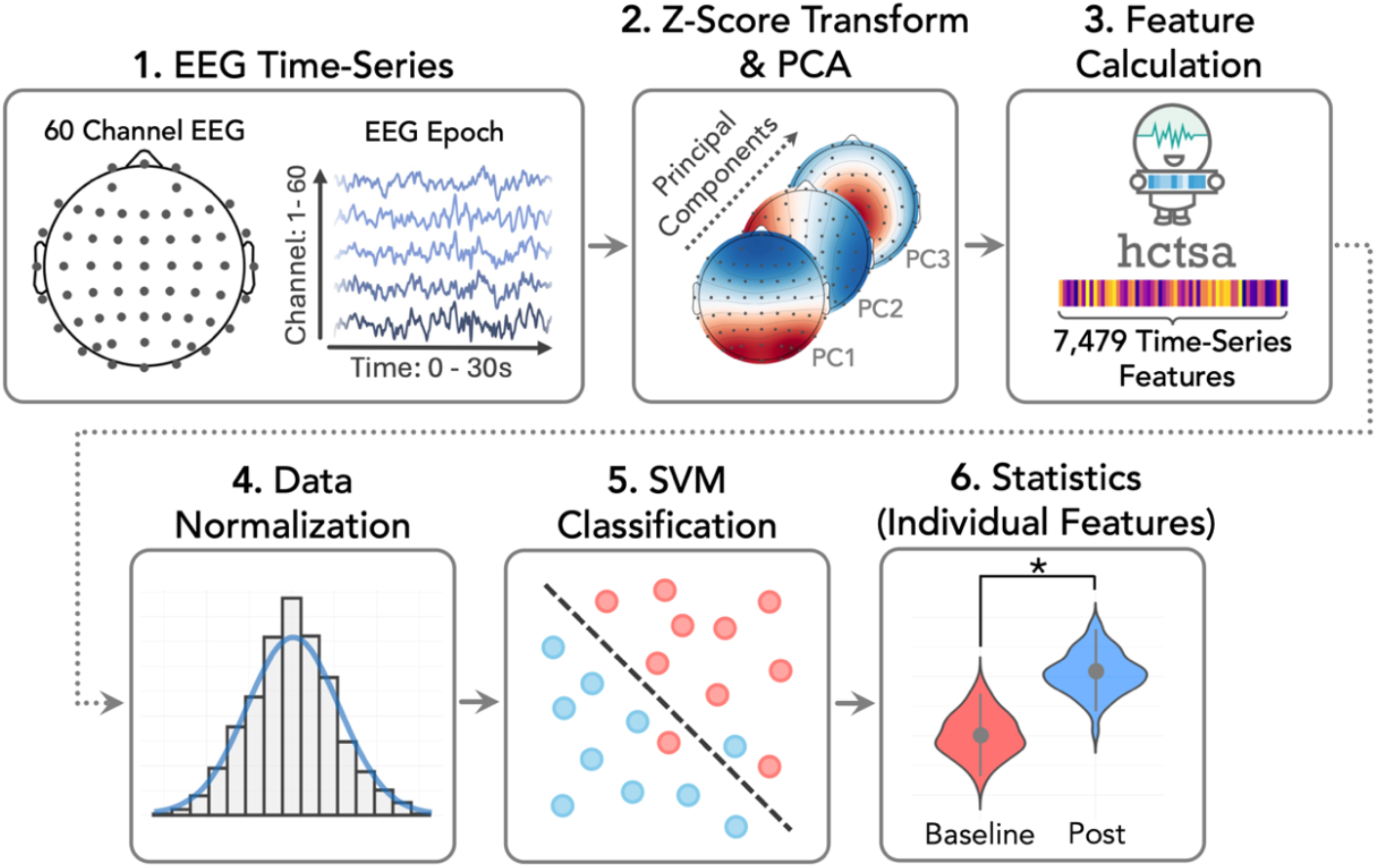
Overview of the data analysis pipeline.

### Statistical Analysis

Performance of each SVM classifier was evaluated against chance using permutation testing (1000 label shuffles [baseline vs post]) to generate a null distribution of mean balanced accuracies. For each PC, *p*-values were obtained by comparing the true balanced accuracy to these shuffles. Outcome *p*-values were FDR-corrected across the 3 PCs [38]. To identify features differentiating pre- and post-treatment EEG data, Mann-Whitney U-tests compared baseline to post conditions for each feature (with statistical significance tested by comparison to Mann-Whitney U-test statistics following 1000 label shuffles) within each PC, with FDR correction across features per PC [39]. Finally, as an exploratory step, for each PC we extracted clusters of related features, then selected two representative top-performing features from each cluster. We then extracted baseline values for these features and tested their combined ability to predict treatment response (>50% HDRS reduction) using a linear SVM (total 18 features). Individual receiver operating characteristic (ROC) analyses (via pROC in R, and SPSS to obtain *p*-values; FDR-corrected) were also run to identify which individual features carried the strongest predictive signal [40].

## Results

The top three PCs explained 78.15% of the total variance in the dataset. Topographic plots and accompanying power spectra for each PC are presented in Figure 2A. Across all three PCs, the spectra show a dominant alpha rhythm; however, as can be seen in the accompanying topographic plots, the electrodes that contributed the strongest weightings to each PC differed substantially, with weightings similar to those reported previously in other studies [25, 33]. SVM classifiers showed above chance classification accuracy of baseline and post-treatment datasets for each of the PCs (all p_FDR_<.001; Table 2). The average null-model classification accuracy from the 1000 null permutations was centred at chance, indicating our approach did not lead to overfitting [25, 33].

**Table 2.** Classification accuracy with accompanying p-values for the top three PCs.

| Principal Component | Variance Explained (%) | Mean Classification Accuracy (SD) | $p_{\text{FDR}}$ | Null Model Accuracy (SD) |
| --- | --- | --- | --- | --- |
| 1 | 44.79 | 76.19 (25.27) | <0.001 | 49.02 (6.88) |
| 2 | 20.16 | 73.81 (25.27) | <0.001 | 49.02 (6.92) |
| 3 | 13.20 | 75.00 (27.61) | <0.001 | 48.52 (7.15) |

**Figure 2.**
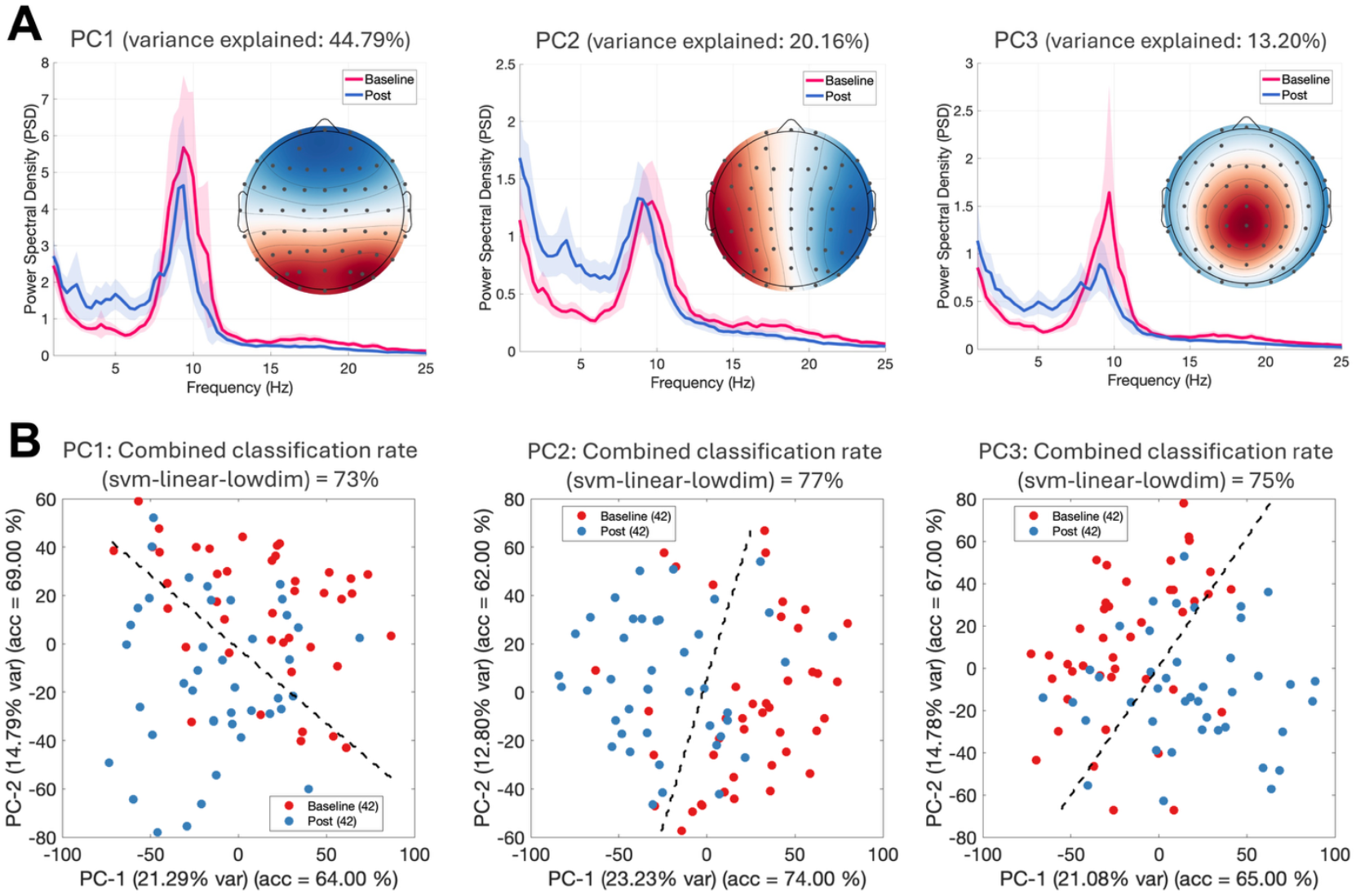
A) Topographic plots of weightings from each electrode that contributed to each of the top three principal components (PCs) with accompanying power spectra. Power spectra were calculated (using the Welch method) across the time-series for each individual participant. Individual spectra were averaged across participants and smoothed using a 3-point centred moving average to aid visualisation. Shaded regions represent 95% confidence intervals around the group mean. B) Low dimensional representations of the normalised data from each PC after *hctsa* feature extraction, using a projection method (PCA) to provide a visual representation of the data in 2-dimensional space using the leading two PCs of the feature-space data matrix. Within these plots, time-series with similar properties are closer in proximity to one-another. Black dotted line denotes the linear classification boundaries. Note, low-dimensional plots serve only as simplified visualisations of the separability of the two classes.

### Top features differentiating baseline and post-treatment neural activity

For PC1, 986 of 7,093 features differed between baseline and post-treatment; for PCs 2 and 3, 1,414 and 1,171 features were significant, respectively (all p_FDR_<.05). To facilitate manageable interpretation, results focussed on the top 40 discriminative features per PC, similar to previous studies [25, 33]. Within each PC, features were clustered by absolute Spearman correlation (|ρ|>0.25), yielding several groups of highly correlated features (Figures 3, 5, 6). Across PCs, two large clusters per PC most strongly separated baseline and post-treatment data, from which we selected a small set of representative, interpretable features for detailed examination.

**Figure 3.**
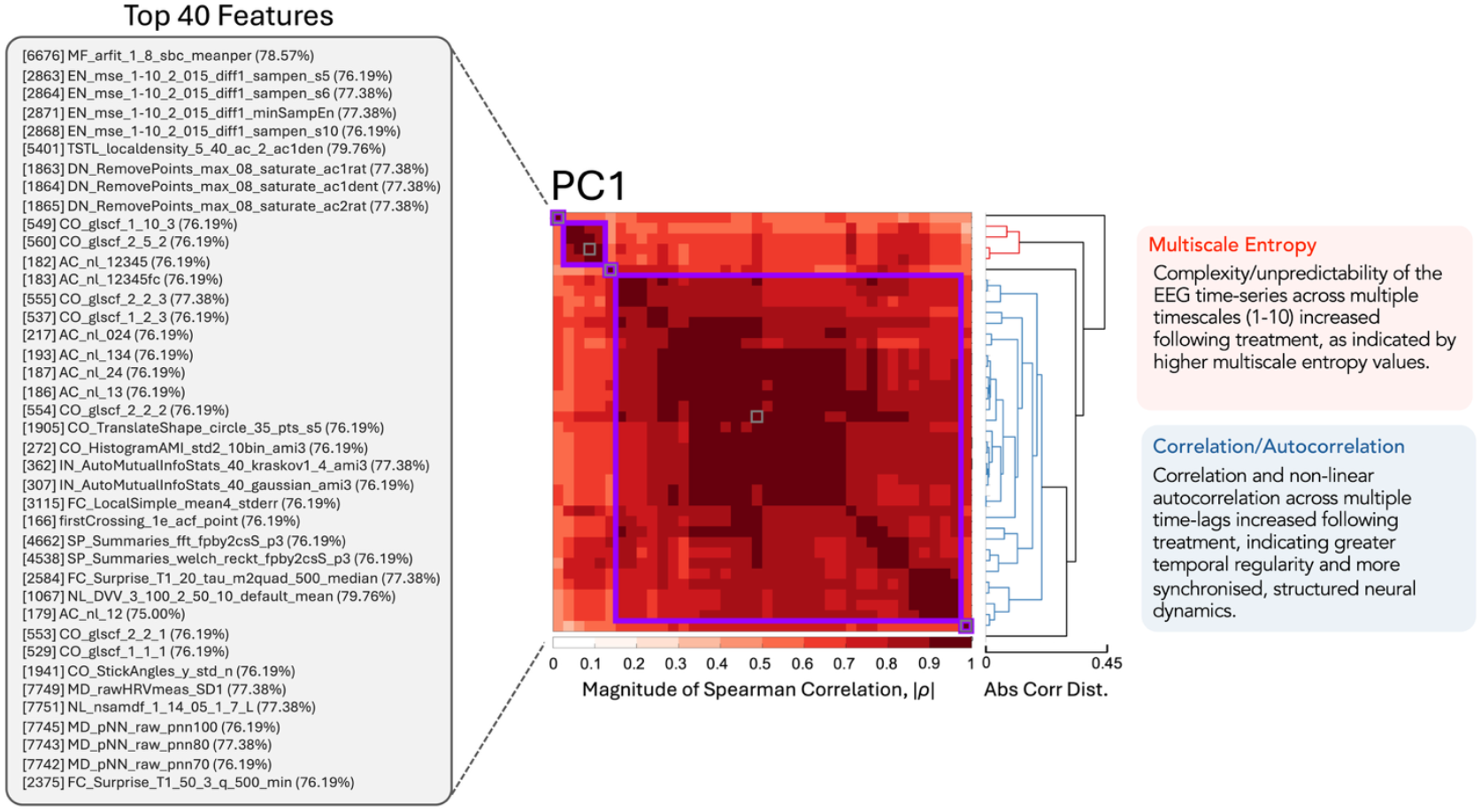
Cluster plots with dendrograms of the top 40 performing features from PC1, which explained 44.79% of the variance in the dataset. Features are clustered using absolute Spearman correlations with a threshold of |ρ| ≥0.25 for cluster formation (clusters indicated with purple borders). Features within the same cluster convey similar information about differences between baseline and post-treatment time-series. Text labels (left) list the feature types from each cluster. Interpretive summaries (right) highlight the dominant feature types from the two largest clusters.

#### PC1

PC1 (posterior dominant; 44.79% variance) showed two clusters of strongly correlated features. One cluster comprised multiscale entropy (MSE) measures from *hctsa* (EN_mse), which estimate signal complexity by coarse-graining the time-series across increasing numbers of consecutive samples (scales) and computing entropy at each scale [41, 42]. Finer scales reflect entropy at faster neural dynamics, whereas coarser scales index slower fluctuations [43]. For the first-differenced series, MSE was higher post-treatment, particularly at scales 5, 6, and 10, indicating that slower neural dynamics became more irregular following treatment. At our sampling rate (160 Hz), these scales correspond to windows of approximately 31, 38 and 63 ms, respectively. Entropy also increased for a measure of the minimum MSE value across scales 1-10 (EN_mse_1-10_2_015_diff1_minSampEn), indicating that convulsive therapy increased the irregularity and complexity of slower neural dynamics, with even the most regular temporal structure becoming more unpredictable. These findings therefore suggest a global shift toward richer multiscale signal structure after treatment. Figure 4A plots mean entropy across scales 1-10 for baseline and post-stimulation PC1 data (computed with the mMSE FieldTrip toolbox) [43-45], showing higher entropy at coarser scales post-treatment (full details in Supplement).

**Figure 4.**
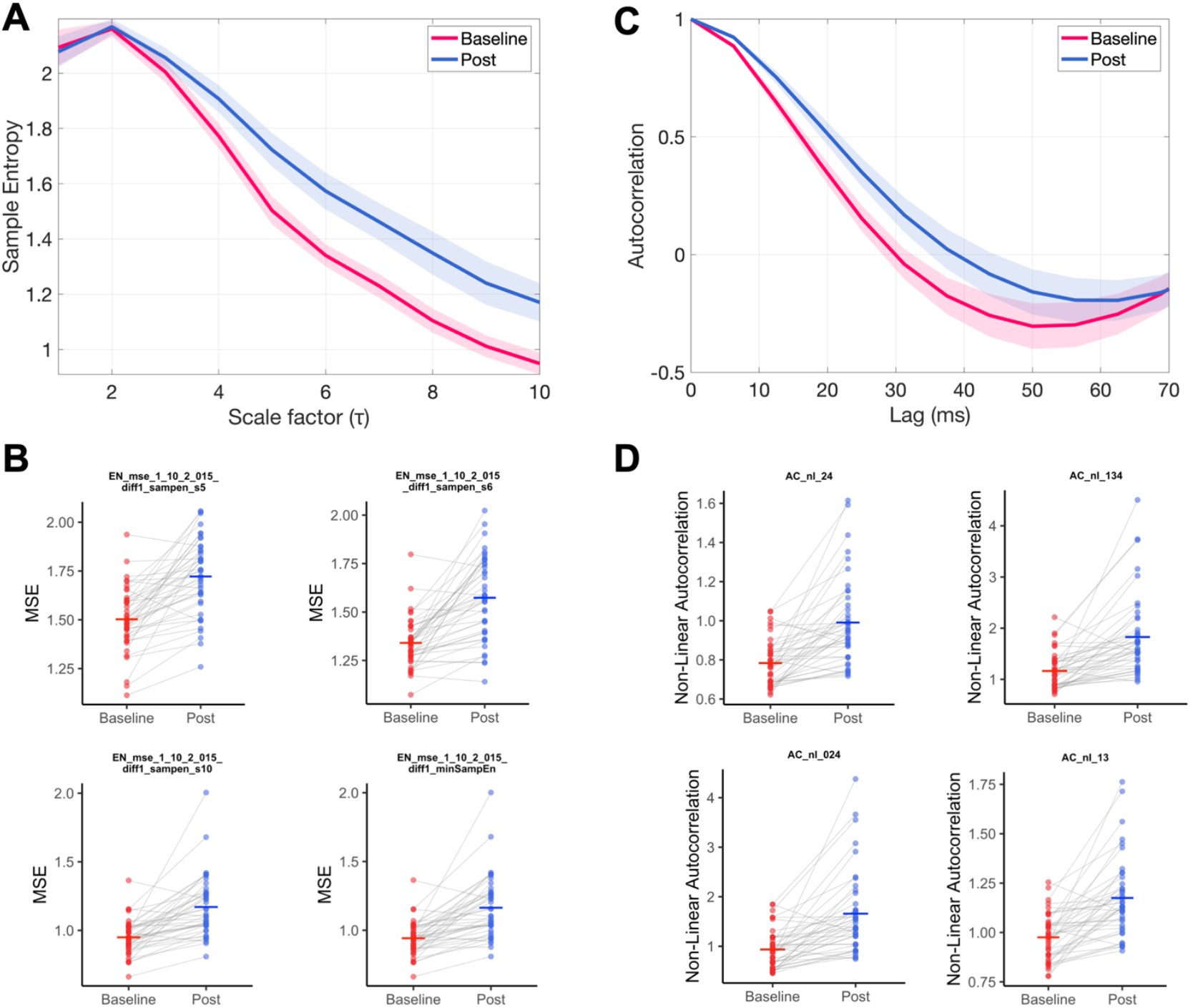
Multiscale entropy and autocorrelation at different timescales or lags. A) Average entropy plotted across scales 1-10 for the baseline and post-stimulation data from PC1 (shading indicates 95% confidence intervals). B) Paired dot plots of four features within the top 40 *hctsa* features from PC1 related to multiscale entropy. Together, these findings highlight a shift towards higher entropy across several scales following convulsive therapy. C) Autocorrelation plot at different lags from PC1, and D) plots for four of the *hctsa* features from PC1 showing greater non-linear autocorrelation post convulsive therapy.

**Figure 5.**
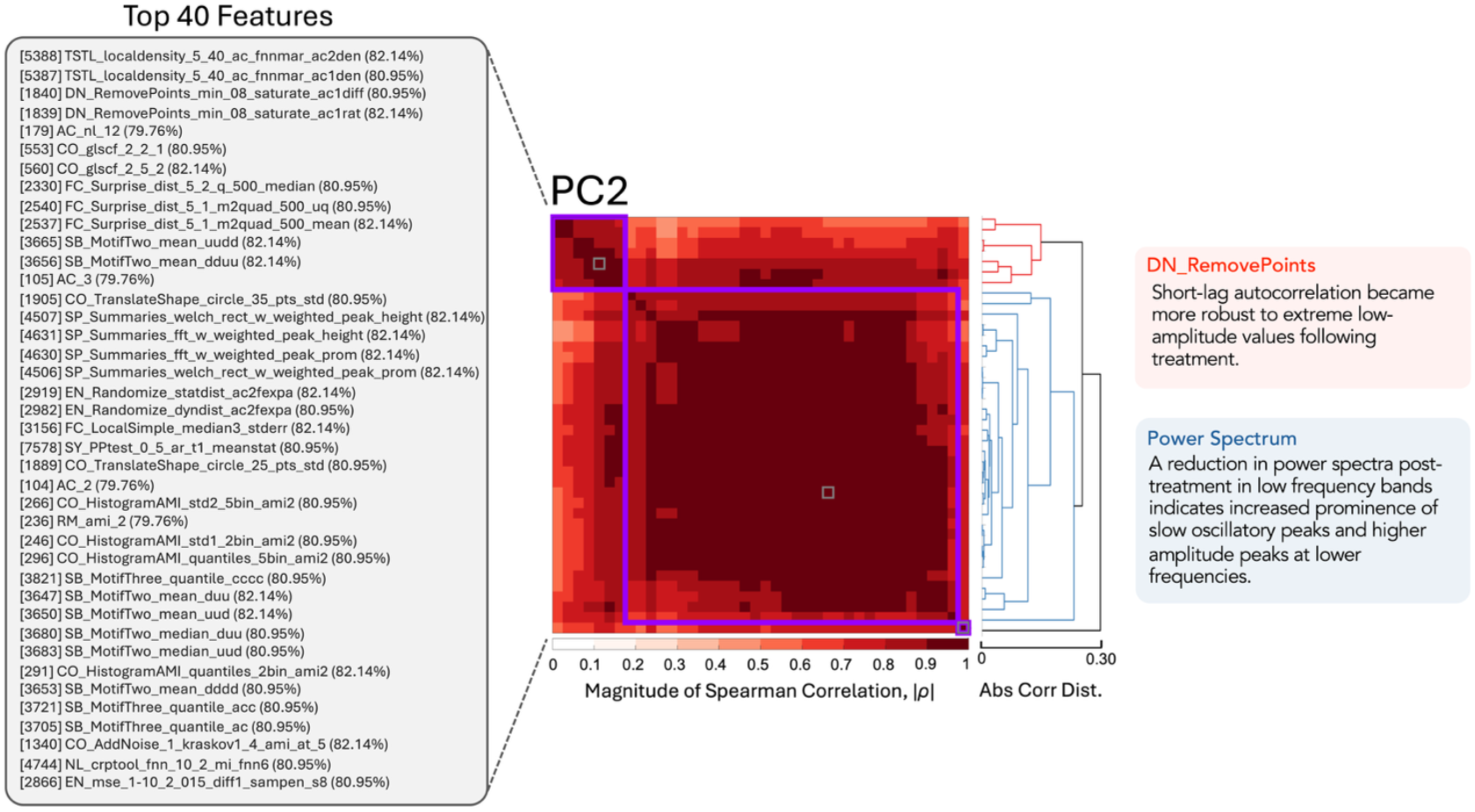
Cluster plots with dendrograms of the top 40 performing features from PC2, which explained 20.16% of the variance in the dataset. Features were clustered using absolute Spearman correlations with a threshold of |ρ| ≥0.25 for cluster formation (clusters indicated with purple borders). Features within the same cluster convey similar information about differences between baseline and post-treatment time-series. Text labels (left) list the feature types from each cluster. Interpretive summaries (right) highlight the dominant feature types from the two largest clusters.

**Figure 6.**
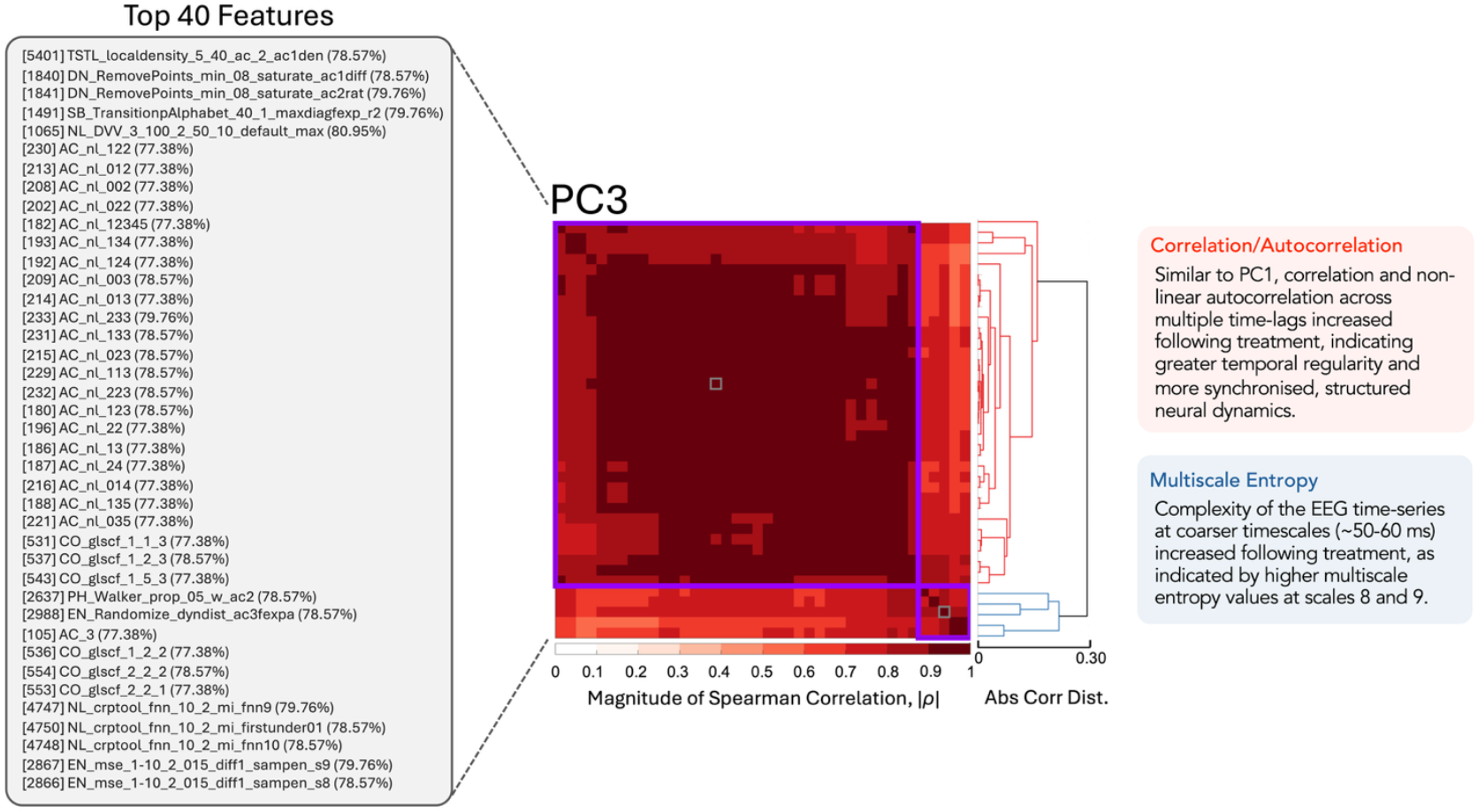
Cluster plots with dendrograms of the top 40 performing features from PC2, which explained 13.20% of the variance in the dataset. Features were clustered using absolute Spearman correlations with a threshold of |ρ| ≥0.25 for cluster formation (clusters indicated with purple borders). Features within the same cluster convey similar information about differences between baseline and post-treatment time-series. Text labels (left) list the feature types from each cluster. Interpretive summaries (right) highlight the dominant feature types from the two largest clusters.

The second PC1 cluster (blue dendrogram in Figure 3) contained most of the remaining top 40 features, predominantly indexing correlation and nonlinear autocorrelation. Non-linear autocorrelation quantifies dependence between a time-series and its lagged versions. These features were consistently higher post-treatment across several short-lag combinations (e.g., AC_nl_024, AC_nl_134, AC_nl_24, AC_nl_13, AC_nl_12, AC_nl_12345), where the numbers in the feature name represent specific lag values, for example, ‘AC_nl_024’ calculates non-linear autocorrelation using lags of 0, 2, and 4 samples (with each lag corresponding to 6.26 ms, given our 160 Hz sampling rate). This pattern indicates stronger short-range regularity and predictability, with enhanced nonlinear structure in the EEG signal at finer time-scales after convulsive therapy (additional illustrative features shown in Supplemental Figure S3). Together, these findings suggest that convulsive therapy increased the short-lag temporal organisation of brain activity, a result that complements the observed entropy changes at broader scales (and previous findings of increased power at lower frequencies), while also highlighting that purely frequency-based features are perhaps not as informative as these more complicated features. To visualise the temporal dependencies underlying these results, we plotted the linear autocorrelation function (ACF) for the same lag sets (e.g., 0-2-4, 1-3-4, 1-5) used by the nonlinear autocorrelation features in *hctsa* (Figure 4C). While these plots reflect linear autocorrelations (rather than the non-linear autocorrelations provided by the higher performing features), they provide an interpretable analogue of the short-lag dependencies that were stronger following treatment.

#### PC 2

For PC2, features related to how time-series properties change as points are removed or their values are saturated (DN_RemovePoints; cluster 1) and the Fourier spectrum (cluster 2) were prominent (Figure 5). DN_RemovePoints features test how fragile the signal is by deleting or flattening small parts of the time series and quantifying how much its overall properties change [22, 24]. Specifically, the two features identified here examine the robustness of lag-1 autocorrelation to distortion of extreme values by saturating the lowest 8% of the signal (i.e., flattening these values to the lowest values). Results indicate that, following treatment, the lag-1 autocorrelation became more stable to this manipulation, indicated by a higher ratio (i.e., ‘ac1rat’) as well as a smaller absolute difference (i.e., ‘ac1diff’) between the original and perturbed signals. Together, these findings suggest that short-lag temporal dependencies in the EEG signal became less influenced by rare low-amplitude events following treatment.

Features related to the Fourier spectrum (e.g., the ‘SP_Summaries’ family of features) were prominent in cluster 2 (highlighted in blue in the dendrogram of Figure 5). Fourier-based features summarised properties of the signal’s spectral composition. Post-treatment, lower values were observed for measures of the weighted height and prominence of the dominant frequency peaks, suggesting that, after treatment, the EEG spectrum was less dominated by strong narrowband oscillatory peaks. This is consistent with a redistribution of spectral energy toward lower frequencies and a reduction in narrowband rhythmic activity. Together, these features highlight that convulsive therapy altered both the temporal stability of the EEG signal and its spectral composition, with dynamics becoming more stable and activity shifting toward slower frequencies (additional plots of key features related to spectral properties are provided in the Supplemental Materials, Figure S4).

#### PC 3

The results for PC3 were similar to PC1, insofar as PC3 was dominated by features relating to the autocorrelation of the time-series and, to a lesser extent, correlation (the ‘AC_nl’ and ‘CO_glscf’ feature families) in the first cluster (Figure 6). Both sets of features provided values that were higher following treatment, indicating stronger short-lag temporal dependencies in the EEG signal. The AC_nl features capture nonlinear predictability across lags of 1-5 samples (∼6-31 ms at the 160 Hz sampling rate), while the CO_glscf features provide linear summaries of short-range autocorrelation structure. Increases across both feature families therefore suggest that convulsive therapy enhanced the fine-timescale linear and non-linear organisation of neural activity, making the signal more regular and predictable over tens of milliseconds. A second smaller cluster included features related to MSE. Similar to PC1, entropy was increased following treatment, specifically at coarser scales (scales 8 and 9, which reflect the ∼50-60 ms timescale range for the 160 Hz sampling rate used here). Together, these findings indicate that treatment simultaneously increased short-lag predictability (autocorrelation) and coarser-scale complexity (entropy) in neural dynamics.

### SVM Classification and Receiver Operating Characteristic Analysis

We trained a linear SVM on the 18 baseline top-performing features to classify responders versus non-responders using leave-one-out cross-validation, evaluating their potential as pretreatment biomarkers [46, 47]. The classifier achieved a balanced accuracy of 0.69 and an AUC of 0.61, significantly exceeding a label-permuted null distribution (1,000 permutations, p=0.014; Figure 7). These findings indicate that the joint top-performing baseline *hctsa* features contain some predictive information about subsequent treatment response. To further identify which individual features carried the strongest signal, we also performed ROC analyses each of the 18 representative features. ROC curves and full metrics are provided in Supplemental Materials (Figure S5; Table S2). Several features showed moderate discrimination (AUCs ∼0.72-0.74), although none remained significant after FDR correction. The highest AUCs were seen for autocorrelation features in PC2 (e.g., AC_3_105 [AUC=0.74]; AC_2_104 [AUC=0.73]), which were lower at baseline in responders, and non-linear autocorrelation feature and spectral peak measures in PC3 (AC_nl_003_209 [AUC=0.72]; SP_Summaries peak height/prominence [both AUC=0.71]), where responders showed higher baseline spectral peaks.

**Figure 7.**
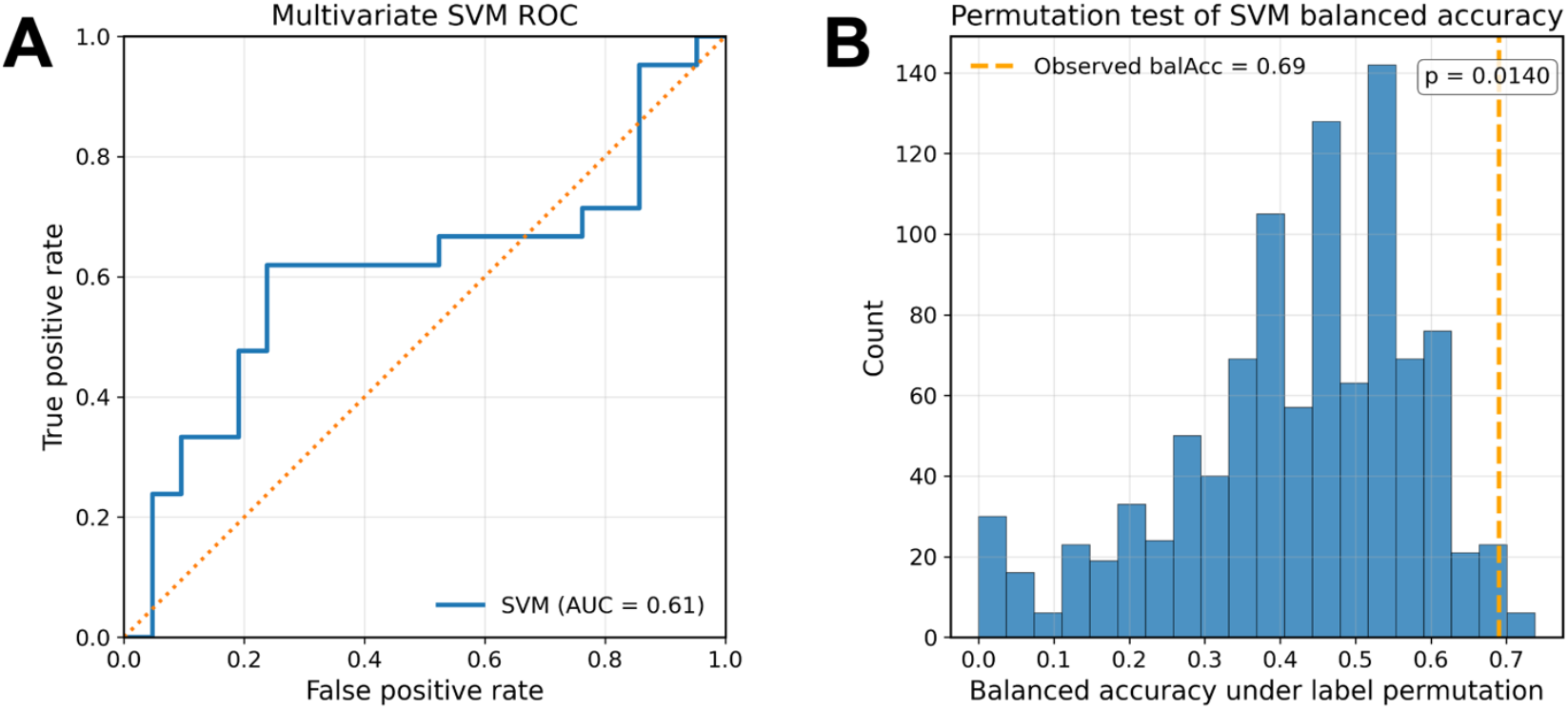
Multivariate support vector machine (SVM) analysis of baseline EEG features for treatment response prediction. A) Receiver operating characteristic (ROC) curve for a linear SVM trained to classify responders versus non-responders using 18 baseline EEG features (top two features per *hctsa* cluster across PCs). Performance was evaluated with leave-one-out cross validation, yielding an area under the curve (AUC) of 0.61. The dashed line indicates chance level (AUC=0.50). B) Null distribution of balanced accuracy obtained by repeating the same leave-one-out analysis after randomly permuting responder labels 1,000 times. The vertical line marks the observed balanced accuracy of 0.69. The permutation test indicated that classifier performance was modest but significantly above chance (p=0.014), consistent with a multivariate predictive signal in the combined baseline feature set.

## Discussion

We utilised a data-driven massive time-series feature extraction approach to examine whether dynamical neural activity patterns can meaningfully distinguish between resting-state EEG recordings taken before and following a treatment course of convulsive therapy. All top three PCs (which accounted for 78.15% of the total variance in the data) demonstrated dynamic patterns that distinctly differentiated baseline from post-treatment recordings with classification accuracies that ranged between 73.8-76.2%. The findings highlight robust alterations in brain dynamics following convulsive therapies for MDD, broadly aligning with previous cross-modal research findings reporting functional brain changes following these interventions [48-50].

Across the top three PCs analysed, between 986-1,414 features significantly differentiated baseline from post-stimulation time-series (after FDR correction for >7,000 comparisons). Examination of these features highlighted changes in signal complexity and predictability (MSE), self-similarity (linear and non-linear autocorrelation), and spectral properties, among many others. A major strength of the massive time-series approach is its capacity to potentially uncover features that have not been previously empirically examined in neuromodulation research or have received only limited investigation. Notably, MSE features appeared consistently within the top 40 examined features for all PCs (Figures 3, 5, and 6), with medium-to-coarse scale entropy increased after treatment, indicating greater signal complexity at these scales.

Few studies have examined EEG-based entropy after convulsive therapy. Our results partly accord with a case report in autism showing increased low-scale (and reduced high-scale) MSE after ECT [51], and with a depression case series showing reduced fine-scale entropy, particularly at finer scales (e.g., scales 1-5) [52]. Another study that used a sample of participants overlapping with this one [28] also reported reductions in complexity at fine timescales, but only in responders to ECT or MST. Together with previous findings, our data indicate that MSE features are likely prominent and discriminative markers of pre-versus post-treatment neural dynamics when analysed within a feature rich framework such as *hctsa*. The fact that the strongest effects occurred at medium-to-coarse scales further suggests that convulsive therapies might preferentially modify slower, integrative neural processes rather than only fast, local dynamics [16] consistent with proposals that therapeutic seizures drive large scale network reorganisation and enhance flexible communication across distributed circuits [14, 18, 53].

Another theme observed across the three PCs was that features indexing correlation and autocorrelation effectively discriminated between pre- and post-treatment timepoints (e.g., CO_glscf and AC_nl, which quantify linear and non-linear time-lagged dependencies). These EEG characteristics have received little attention in convulsive therapy research. Autocorrelation was generally higher post-treatment, indicating greater self-similarity and predictability, possibly reflecting more stable oscillatory activity and greater large-scale coordination, most evident at short lags (e.g., AC_nl_12, AC_nl_13 [PC1]; AC_2, AC_3, CO_glscf_2_5_2 [PC2]; AC_nl_002 [PC3]). Such changes may partly reflect a shift in spectral content, with reduced high-frequency and increased low-frequency power enhancing short-lag autocorrelation. In contrast, MSE features commonly emerged at medium-to-coarse scales (e.g., s5-s10), indicating increased complexity over slower timescales. Together, these findings suggest that convulsive therapy alters higher-order temporal structure beyond what is captured by standard power-spectrum measures, thus highlighting the value of *hctsa* for quantifying more nuanced treatment-related neural dynamics.

We know from past research that convulsive therapies can lead to enhanced high amplitude slow-wave (e.g., theta and delta) activity on the EEG record, often also accompanied by reduced higher frequency (beta, gamma) activity, which is likely related to residual post-ictal phenomena as a result of the induced seizure [8]. This has been reported by others [8, 54, 55], as well as in the current cohort when analysing the Fourier spectra following both ECT [14] and MST [17]. This pattern is also reflected in our data-driven analysis through features related to the power spectrum, i.e., ‘SP_Summaries_welch’ and ‘SP_Summaries_fft’, which measure spectral properties of the data after transforming the signal to the frequency domain using an FFT or Welch approach, respectively. These features were observed in the top 40 features for PC1 (2 features) and PC2 (4 features), suggesting that oscillatory peaks were both more prominent, and of a higher amplitude at lower frequencies following treatment (specific examples in Supplemental Figure S3).

Our exploratory ROC analyses suggested moderate discrimination between responders and non-responders for several baseline features, with linear and non-linear autocorrelation measures performing particularly well. Although no single feature survived FDR-correction, the convergence of autocorrelation-based measures across the pre-versus post-treatment *hctsa* analysis, and within the ROC treatment response classification results, suggests that changes in the predictability or self-similarity of neural dynamics may represent a promising and previously under-recognised marker of therapeutic response. Further, when we combined the 18 top-performing baseline features in a multivariate model, a linear SVM achieved modest but statistically reliable discrimination of responders versus non-responders (p=0.014). This suggests that baseline *hctsa* features might carry predictive information primarily as a distributed pattern across complementary dynamics, rather than through any single feature in isolation.

More broadly, our findings help address the longstanding disconnect between the high efficacy of convulsive therapies in treatment-resistant MDD, and the limited understanding of their neurobiological mechanisms [7, 56]. Our results point to a shift in the temporal organisation of spontaneous neural activity spanning increased medium-to-coarse scale complexity altered short-lag temporal dependence (linear and non-linear autocorrelation), and spectral slowing. A possible interpretation is that convulsive therapy induces large-scale neural perturbations that transiently disrupt maladaptive network dynamics enabling plastic reconfiguration across distributed circuits consistent with evidence of connectivity and plasticity-related changes following ECT and MST [14, 17, 18, 50, 57, 58]. Notably, the shift towards longer time-scales appears broadly consistent with hierarchical temporal organisation of the cortex, whereby association regions (including frontal cortex) integrate information over broader temporal windows than sensory regions, supporting contextual integration and top-down modulation of lower-level processing [59, 60]. While speculative, this could provide a potential mechanistic bridge between the changes in neural dynamics we observed to be associated with convulsive therapies and symptom improvement, with a broader temporal integration in neural processing perhaps enabling enhanced contextual regulation of mood, greater flexibility of thought, and more effective inhibition of narrow, reactive emotional responses [59, 61, 62]. Future work could extend this framework to derive robust multi-feature profiles predictive of response, supporting development of mechanistically grounded biomarkers for precision psychiatry [63, 64].

### Limitations and Future Research

Our study has several limitations. First, ECT and MST were pooled to increase power, precluding direct comparisons, so findings reflect convulsive therapy broadly, rather than modality-specific effects. We also did not test whether the full *hctsa* feature set could distinguish clinical responders from non-responders, given the limited sample size. Instead, we trained an SVM on a limited set of top-performing representative baseline features and evaluated performance with cross-validation. Although performance exceeded a permutation-based null, estimates require replication in larger cohorts. Second, as *hctsa* requires univariate time-series, we applied PCA and analysed the first three components, capturing the majority of variance, but sacrificing spatial information across electrodes. Third, as the MST and ECT trials lacked a placebo comparison group, within-subject effects could not be adequately controlled for. Many participants were also taking medications that may have influenced brain activity. Medication regimens, however, were stable across recordings, so they cannot explain within-subject pre-post differences. Finally, only baseline and relatively immediate post-treatment EEG were available, limiting inferences about the durability and longer-term evolution of neural changes.

## Conclusions

Our findings demonstrate that a broad spectrum of time-series features derived from resting-state EEG can reliably distinguish post-convulsive therapy neural dynamics from baseline activity. This data-driven framework offers a powerful and objective means of characterising how therapeutic brain stimulation reshapes neural activity, providing new insights into the mechanisms underlying treatment effects. Future research extending this to other therapeutic neurostimulation interventions holds substantial potential.

## Data Availability

Data presented in the study is not available at this time.

## Funding Information

In the last 3 years PBF has received equipment for research from Neurosoft and Nexstim. He has served on a scientific advisory board for Magstim and received speaker fees from Otsuka. He has also acted as a founder and board member for TMS Clinics Australia and Resonance Therapeutics. PBF is supported by a National Health and Medical Research Council of Australia Investigator grant (1193596). ZJD has received research and equipment in-kind support for an investigator-initiated study through Brainsway Inc and Magventure Inc and industry-initiated trials through Magnus Inc. He also currently serves on the scientific advisory board for Brainsway Inc. His work has been supported by the National Institutes of Mental Health (NIMH), the Canadian Institutes of Health Research (CIHR), Brain Canada and the Temerty Family, Grant and Kreutzcamp Family Foundations. DMB receives research support from CIHR, NIMH, Wellcome Trust, Brain Canada and the Temerty Family through the CAMH Foundation and the Campbell Family Research Institute, has received research grant support and in-kind equipment support for an investigator-initiated study from Brainsway Ltd, was the site principal investigator for three sponsor-initiated studies for Brainsway Ltd, has received in-kind equipment support from Magventure for investigator-initiated studies, has received medication supplies for an investigator-initiated trial from Indivior, is a scientific advisor for Sooma Medical and is a member of the Clinical Standards Committee of the Clinical TMS Society (unpaid). ATH, KG, JAGL, RZ, and NWB have nothing to declare.

## Notes

### Clinical Trial

NCT01596608

### Funding Statement

This study did not receive any funding.

### Author Declarations

Ethics Committee of the Center for Addiction and Mental Health gave ethical approval for the study in accordance with the declaration of Helsinki.

