## Supplementary Material for "Uncovering Neural Signatures of Convulsive Therapy in Depression Using Massive EEG Time-Series Feature Extraction"

### Supplementary Methods and Materials

*Convulsive Therapy*

Electroconvulsive therapy (ECT) was administered with a brief-pulse square-wave device (MECTA Corporation, Oswego, OR), 2-3 times per week. Thirteen subjects began with right unilateral ultra-brief ECT (0.3 msec pulse width), and one subject with bitemporal ECT, based on physician and patient preference. Electrode placement followed American Psychiatric Association guidelines [1]. The bitemporal montage placed the electrode centre 2-3 cm above the midpoint between the outer canthus of the eye and the external auditory canal [2]. For right unilateral ECT, one electrode was placed using the same coordinate as the bitemporal montage, with the other electrode placed 2–3 cm to the right of the vertex [3]. Five unilateral subjects later switched to bitemporal ECT due to insufficient clinical response. Anaesthesia was induced using methohexital and succinylcholine for muscle relaxation. Treatment completion was determined by clinical factors, treatment response, the patient's preference to discontinue, and the clinical judgment by the treating physician. A detailed procedure can be found in Voineskos et al. [4].

Magnetic seizure therapy (MST) was delivered using a MagPro MST stimulator (Magvenure, Denmark) with a Twin Coil-XS, applied bilaterally with the centre of each coil over prefrontal sites approximating F3 and F4 (10-20 system). With this positioning the maximum E-field was directed over the dorsomedial prefrontal cortex. MST was administered 2-3 times per week until either remission (HDRS-24 score ≤ 10 and ≥60% symptom reduction on two consecutive ratings) or a maximum of 24 treatments. All sessions were conducted under general anaesthesia (methohexital sodium or methohexital + remifentanil) with neuromuscular blockade (succinylcholine). A detailed procedure is described in Daskalakis et al. [5].

*EEG Data Acquisition and Pre-Processing*

Eyes-closed resting-state electroencephalography (EEG) data were recorded for 10 minutes using a 64-channel Neuroscan *Quik-cap* connected to a SynAmps$^2$ amplifier (Compumedics, USA). Recordings used a sampling rate of either 1 KHz or 10 KHz (with 10 KHz files downsampled to 1 KHz prior to pre-processing) and online low- and high-pass filters at 1 KHz and 0.05 Hz, respectively. The online reference electrode was positioned at the vertex (between CPz and Cz), and the ground was positioned on the midline, just posterior to Fz. Baseline EEG recordings were performed in the week immediately prior to commencement of convulsive therapy. For the ECT dataset, post-treatment EEG recordings were collected within two days following the final treatment session; while for MST, data were collected on average 3.81 (SD = 3.86) days following the final treatment session.

All EEG data were cleaned using the automated RELAX (Reduction of Electroencephalographic Artefacts) pipeline, version 2.0.0 [6-8] in MATLAB. RELAX uses empirical methods to identify and reduce artefacts in the data, including wavelet-enhanced independent component analysis (wICA) targeted to artefact periods within the data [8]. Full details of the RELAX pipeline can be found in Bailey et al. [6, 7]. In brief, data were bandpass filtered (0.5-80 Hz; acausal Butterworth filter) and notch filtered (57-63 Hz) to eliminate electrical line noise. Noisy channels were removed through a multi-step process incorporating the 'findNoisyChannels' function from the PREP pipeline [9] and RELAX default outlier detection functions. Extremely noisy periods of the data were detected and removed using the same RELAX default outlier detection functions (with the default "moderate" rejection settings from RELAX v2.0.0: [8]). Artefact components were identified using the IClabel machine learning algorithm [10] and icablinkmetrics [11]. Targeted wICA was applied to clean artefacts across specific artefact periods (for ocular artefacts) and frequencies (for muscle components), which can help to preserve the neural signal, while still effectively removing artefacts. Specifically, targeted wICA weights artefact reduction for eye-movement components before applying it within each ICA component, with weights of 1 assigned to periods likely to contain artefact (based on outlier thresholds), weights of 0 outside those periods, and a progressive increase in weighting from 0 to 1 over the 200 ms surrounding each artefact [12].

Finally, data were segmented into 30-second epochs, with any epochs showing remaining artefacts removed using RELAX's default settings applied (with the exception of absolute voltage amplitude being 120 μV to avoid rejection of high-amplitude alpha activity and/or treatment-induced slow-wave activity). 30-second epochs were chosen as this epoch length has been successfully implemented in previous studies using the *hctsa* approach [13, 14], while also maximising data retention, as most participants had at least one artefact-free epoch available. During this process, three participants were removed from further analysis. One did not have any artifact free baseline or post-treatment 30-second epochs available; another did not have any available post-treatment 30 second epochs; and one participant had excessive noise on their post-treatment EEG record that persisted after cleaning. This resulted in a total of 42 participants (21 ECT, 21 MST) with complete baseline and post-treatment data included in analyses. For all analyses the MST and ECT data were combined to maximise statistical power for the analyses.

The first artefact-free 30-second epoch available in each dataset was then used for all subsequent analyses. The data were down-sampled to 160 Hz to reduce computational burden and focus analyses on frequencies that contain the majority of meaningful EEG variance for time-series feature calculation. Data were then z-score transformed across all electrodes and timepoints separately within each participant's 30s epoch, thus normalising the 60-electrode by 4800-timepoint data [13].This procedure adjusted for overall voltage amplitude differences across participants' EEG data prior to PCA, while preserving the relationships between electrodes. As the EEG data were multivariate (i.e., 60 time-series, one for each electrode) and *hctsa* requires a single univariate time-series, we followed an approach similar to previous studies by applying the linear dimensionality reduction technique, principal component analysis (PCA), to reduce the EEG data to three PCs that captured significant variance in the EEG data. Prior to performing PCA, the time-series data were concatenated in the temporal dimension, and the PCA decomposition was performed on this concatenated time-series, following which the weightings were applied to each individual. This approach ensured that all PCA weights were matched across participants [13, 15]. The z-score transform also prevented potential class imbalances in EEG amplitudes from disproportionately influencing the PCA components towards individuals with higher amplitude signals. Within our data the top 3 PCs explained 78.15% variance in the data and were selected for further analysis.

*Statistical Analysis*

To enable statistical testing of how the SVM classifier's performance compared to chance, we implemented a permutation approach which involved calculating the probability of obtaining the observed mean balanced accuracy in relation to a null distribution with 1000 null samples, whereby each null sample involved training an SVM classifier on data using shuffled labels (i.e., baseline, post) [16]. This provides a test of the likelihood of observing the classification performance observed using the true labels, in the context of a distribution of classification accuracies obtained from randomised labels. False discovery rate (FDR) corrections were then further used to control for models run across each of the 3 PCs [17]. Finally, we assessed the capacity of individual features to discriminate between baseline and post-stimulation data, identifying the individual features associated with accurate predictions. This was implemented by performing non-parametric Mann-Whitney U-tests on each feature independently, within each PC, and comparing the Mann-Whitney U test statistics obtained from tests on the real data to a distribution of Mann-Whitney U test statistics obtained from 1000 shuffles of the labels. This approach enabled us to infer which types of dynamical neural changes captured by a given PC might have contributed to classification performance. To correct for multiple comparisons, FDR corrections were applied across all features within each PC. As a final exploratory step, for each PC, we selected several top performing representative features from each feature cluster, and extracted these features from the baseline EEG data, then tested whether they could predict participant treatment responder status (with responders classified as participants with >50% reductions in HDRS scores following completion of treatment) using receiver operating characteristic analyses. These analyses were performed in R using the pROC package [18] and also in SPSS (to obtain p-values), which were FDR-corrected.

*Generation of example entropy and autocorrelation plots used in Figure 4*

#### *Visualisation of multiscale entropy*

To better illustrate the changes in multiscale entropy (MSE) identified by *hctsa* for PC1, we ran an additional analysis using the mMSE fieldtrip toolbox (https://github.com/LNDG/mMSE) [19-21]. MSE was calculated separately for the Baseline and Post conditions for PC1. Analysis parameters were chosen to align closely with the configuration used by *hctsa* ('EN_mse'). Sample entropy was estimated with an embedding dimension of m = 2 and a similarity tolerance of r = 0.15 x SD. The full trial length was used as the analysis window (timwin), centred on the median time point (toi). MSE was computed over scales 1-10 using the point averaging coarse-graining method, with a fixed similarity bound applied across scales (recompute_r = per_toi). In the point averaging coarse-graining procedure, the original time series is rescaled by averaging non-overlapping windows of data points. For example, at scale 2, successive pairs of samples are averaged to form a new, shorter time series; at scale 5, groups of five consecutive samples are averaged, and so on. Sample entropy is then calculated on each coarse-grained series. This method corresponds to the classic implementation of MSE introduced by Costa et al. [22] and closely matches the approach used in *hctsa*. The results of this analysis can be seen in Figure 5 within the main manuscript. Briefly, the multiscale entropy curves show that, while both the baseline and post-treatment conditions followed the expected pattern of decreasing entropy with increasing scale factor, the post-treatment condition consistently demonstrated higher entropy values, especially from approximately a scale factor ($\tau$) of 3 through to $\tau$=10. This suggests that treatment was associated with an increase in signal complexity and unpredictability across medium and coarser timescales.

#### *Visualisation of autocorrelation features*

Nonlinear autocorrelation features identified by *hctsa* (e.g., AC_nl_024, AC_nl_134, AC_nl_12345) are computed as m-point autocorrelations of the z-scored time series at the specified short lags, i.e., the mean product of the signal with its lagged copies. Larger values indicate stronger higher-order temporal dependence across those lags, beyond what is captured by the standard two-point autocorrelation. To aid interpretation, we additionally plotted the linear autocorrelation function (ACF) for the same lag sets used in the *hctsa* features. The ACF was computed using mean-removed signals, normalised such that ACF(0) = 1, and averaged across participants within the Baseline and Post conditions. While these plots reflect linear autocorrelation rather than the nonlinear statistic used by *hctsa*, they provide an intuitive analogue of the short-lag temporal dependencies (1-5 samples; ~6-31 ms at 160 Hz) that were found to increase following treatment.

**Table S1:** List of Top 40 Features for each Principal Component

| | Feature Number | Feature Name | Time-Series Analysis Class (keyword) | Balanced Accuracy |
|---|---|---|---|---|
| PC1 | | | | |
| | [1067] | NL_DVV_3_100_2_50_10_default_mean | (delayVectorVariance) | 79.76% |
| | [5401] | TSTL_localdensity_5_40_ac_2_ac1den | (nonlinear,tstool) | 79.76% |
| | [6676] | MF_arfit_1_8_sbc_meanper | (modelfit,arfit) | 78.57% |
| | [362] | IN_AutoMutualInfoStats_40_kraskov1_4_ami3 | (information,correlation,AMI) | 77.38% |
| | [555] | CO_glscf_2_2_3 | (correlation,glscf) | 77.38% |
| | [1863] | DN_RemovePoints_max_08_saturate_ac1rat | (correlation,outliers) | 77.38% |
| | [1864] | DN_RemovePoints_max_08_saturate_ac1diff | (correlation,outliers) | 77.38% |
| | [1865] | DN_RemovePoints_max_08_saturate_ac2rat | (correlation,outliers) | 77.38% |
| | [2584] | FC_Surprise_T1_20_tau_m2quad_500_median | (information,symbolic) | 77.38% |
| | [2864] | EN_mse_1-10_2_015_diff1_sampen_s6 | (entropy) | 77.38% |
| | [2871] | EN_mse_1-10_2_015_diff1_minSampEn10_2_015_diff1_minSampEn | (entropy) | 77.38% |
| | [7743] | MD_pNN_raw_pnn80 | (medical,raw,spreadDependent) | 77.38% |
| | [7749] | MD_rawHRVmeas_SD1 | (medical,raw,spreadDependent) | 77.38% |
| | [7751] | NL_nsamdf_1_14_05_1_7_L | (nonlinear,raw,spreadDependent) | 77.38% |
| | [166] | firstCrossing_1e_acf_point | (correlation,timescale) | 76.19% |
| | [182] | AC_nl_12345 | (correlation,nonlinearautocorr) | 76.19% |
| | [183] | AC_nl_123456 | (correlation,nonlinearautocorr) | 76.19% |
| | [186] | AC_nl_13 | (correlation,nonlinearautocorr) | 76.19% |
| | [187] | AC_nl_24 | (correlation,nonlinearautocorr) | 76.19% |
| | [193] | AC_nl_134 | (correlation,nonlinearautocorr) | 76.19% |
| | [217] | AC_nl_024 | (correlation,nonlinearautocorr) | 76.19% |
| | [272] | CO_HistogramAMI_std2_10bin_ami3 | (information,correlation,AMI) | 76.19% |
| | [307] | IN_AutoMutualInfoStats_40_gaussian_ami3 | (information,correlation,AMI) | 76.19% |
| | [529] | CO_glscf_1_1_1 | (correlation,glscf) | 76.19% |
| | [537] | CO_glscf_1_2_3 | (correlation,glscf) | 76.19% |

| | | | |
|---|---|---|---|
| [549] | CO_glscf_1_10_3 | (correlation,glscf) | 76.19% |
| [553] | CO_glscf_2_2_1 | (correlation,glscf) | 76.19% |
| [554] | CO_glscf_2_2_2 | (correlation,glscf) | 76.19% |
| [560] | CO_glscf_2_5_2 | (correlation,glscf) | 76.19% |
| [1905] | CO_TranslateShape_circle_35_pts_std | (correlation) | 76.19% |
| [1941] | CO_StickAngles_y_std_n | (correlation) | 76.19% |
| [2375] | FC_Surprise_T1_50_3_q_500_min | (information,symbolic) | 76.19% |
| [2863] | EN_mse_1-10_2_015_diff1_sampen_s5 | (entropy) | 76.19% |
| [2868] | EN_mse_1-10_2_015_diff1_sampen_s10 | (entropy) | 76.19% |
| [3115] | FC_LocalSimple_mean4_stderr | (forecasting) | 76.19% |
| [4538] | SP_Summaries_welch_rect_fpoly2csS_p3 | (FourierSpectrum) | 76.19% |
| [4662] | SP_Summaries_fft_fpoly2csS_p3 | (FourierSpectrum) | 76.19% |
| [7742] | MD_pNN_raw_pnn70 | (medical,raw,spreadDependent) | 76.19% |
| [7745] | MD_pNN_raw_pnn100 | (medical,raw,spreadDependent) | 76.19% |
| [179] | AC_nl_12 | (correlation,nonlinearautocorr) | 75.00% |

PC2

| | | | |
|---|---|---|---|
| [291] | CO_HistogramAMI_quantiles_2bin_ami2 | (information,correlation,AMI) | 82.14% |
| [560] | CO_glscf_2_5_2 | (correlation,glscf) | 82.14% |
| [1340] | CO_AddNoise_1_kraskov1_4_ami_at_5 | (correlation,AMI,entropy) | 82.14% |
| [1839] | DN_RemovePoints_min_08_saturate_ac1rat | (correlation,outliers) | 82.14% |
| [2537] | FC_Surprise_dist_5_1_m2quad_500_mean | (information,symbolic) | 82.14% |
| [2919] | EN_Randomize_statdist_ac2fexpa | (entropy,slow) | 82.14% |
| [3156] | FC_LocalSimple_median3_stderr | (forecasting) | 82.14% |
| [3647] | SB_MotifTwo_mean_duu | (symbolic,motifs) | 82.14% |
| [3650] | SB_MotifTwo_mean_uud | (symbolic,motifs) | 82.14% |
| [3656] | SB_MotifTwo_mean_dduu | (symbolic,motifs) | 82.14% |
| [3665] | SB_MotifTwo_mean_uudd | (symbolic,motifs) | 82.14% |
| [4506] | SP_Summaries_welch_rect_w_weighted_peak_prom | (FourierSpectrum) | 82.14% |
| [4507] | SP_Summaries_welch_rect_w_weighted_peak_height | (FourierSpectrum) | 82.14% |

| | | | |
|---|---|---|---|
| [4630] | SP_Summaries_fft_w_weighted_peak_prom | (FourierSpectrum) | 82.14% |
| [4631] | SP_Summaries_fft_w_weighted_peak_height | (FourierSpectrum) | 82.14% |
| [5388] | TSTL_localdensity_5_40_ac_fnnmar_ac2den | (nonlinear,tstool) | 82.14% |
| [246] | CO_HistogramAMI_std1_2bin_ami2 | (information,correlation,AMI) | 80.95% |
| [266] | CO_HistogramAMI_std2_5bin_ami2 | (information,correlation,AMI) | 80.95% |
| [296] | CO_HistogramAMI_quantiles_5bin_ami2 | (information,correlation,AMI) | 80.95% |
| [553] | CO_glscf_2_2_1 | (correlation,glscf) | 80.95% |
| [1840] | DN_RemovePoints_min_08_saturate_ac1diff | (correlation,outliers) | 80.95% |
| [1889] | CO_TranslateShape_circle_25_pts_std | (correlation) | 80.95% |
| [1905] | CO_TranslateShape_circle_35_pts_std | (correlation) | 80.95% |
| [2330] | FC_Surprise_dist_5_2_q_500_median | (information,symbolic) | 80.95% |
| [2540] | FC_Surprise_dist_5_1_m2quad_500_uq | (information,symbolic) | 80.95% |
| [2866] | EN_mse_1-10_2_015_diff1_sampen_s8 | (entropy) | 80.95% |
| [2982] | EN_Randomize_dyndist_ac2fexpa | (entropy,slow) | 80.95% |
| [3653] | SB_MotifTwo_mean_dddd | (symbolic,motifs) | 80.95% |
| [3680] | SB_MotifTwo_median_duu | (symbolic,motifs) | 80.95% |
| [3683] | SB_MotifTwo_median_uud | (symbolic,motifs) | 80.95% |
| [3705] | SB_MotifThree_quantile_ac | (symbolic,motifs) | 80.95% |
| [3721] | SB_MotifThree_quantile_acc | (symbolic,motifs) | 80.95% |
| [3821] | SB_MotifThree_quantile_cccc | (symbolic,motifs) | 80.95% |
| [4744] | NL_crptool_fnn_10_2_mi_fnn6 | (nonlinear,dimension,crptool) | 80.95% |
| [5387] | TSTL_localdensity_5_40_ac_fnnmar_ac1den | (nonlinear,tstool) | 80.95% |
| [7578] | SY_PPtest_0_5_ar_t1_meanstat | (unitroot) | 80.95% |
| [104] | AC_2 | (correlation) | 79.76% |
| [105] | AC_3 | (correlation) | 79.76% |
| [179] | AC_nl_12 | (correlation,nonlinearautocorr) | 79.76% |
| [236] | RM_ami_2 | (information,correlation,AMI) | 79.76% |

PC3

| | | | |
|---|---|---|---|
| [1065] | NL_DVV_3_100_2_50_10_default_max | (delayVectorVariance) | 80.95% |

| | | | |
|---|---|---|---|
| [233] | AC_nl_233 | (correlation,nonlinearautocorr) | 79.76% |
| [1491] | SB_TransitionpAlphabet_40_1_maxdiagfexp_r2 | (symbolic,transitionmat) | 79.76% |
| [1841] | DN_RemovePoints_min_08_saturate_ac2rat | (correlation,outliers) | 79.76% |
| [2867] | EN_mse_1-10_2_015_diff1_sampen_s9 | (entropy) | 79.76% |
| [4747] | NL_crptool_fnn_10_2_mi_fnn9 | (nonlinear,dimension,crptool) | 79.76% |
| [180] | AC_nl_123 | (correlation,nonlinearautocorr) | 78.57% |
| [209] | AC_nl_003 | (correlation,nonlinearautocorr) | 78.57% |
| [215] | AC_nl_023 | (correlation,nonlinearautocorr) | 78.57% |
| [229] | AC_nl_113 | (correlation,nonlinearautocorr) | 78.57% |
| [231] | AC_nl_133 | (correlation,nonlinearautocorr) | 78.57% |
| [232] | AC_nl_223 | (correlation,nonlinearautocorr) | 78.57% |
| [537] | CO_glscf_1_2_3 | (correlation,glscf) | 78.57% |
| [554] | CO_glscf_2_2_2 | (correlation,glscf) | 78.57% |
| [1840] | DN_RemovePoints_min_08_saturate_ac1diff | (correlation,outliers) | 78.57% |
| [2637] | PH_Walker_prop_05_w_ac2 | (trend) | 78.57% |
| [2866] | EN_mse_1-10_2_015_diff1_sampen_s8 | (entropy) | 78.57% |
| [2988] | EN_Randomize_dyndist_ac3fexpa | (entropy,slow) | 78.57% |
| [4748] | NL_crptool_fnn_10_2_mi_fnn10 | (nonlinear,dimension,crptool) | 78.57% |
| [4750] | NL_crptool_fnn_10_2_mi_firstunder01 | (nonlinear,dimension,crptool) | 78.57% |
| [5401] | TSTL_localdensity_5_40_ac_2_ac1den | (nonlinear,tstool) | 78.57% |
| [105] | AC_3 | (correlation) | 77.38% |
| [182] | AC_nl_12345 | (correlation,nonlinearautocorr) | 77.38% |
| [186] | AC_nl_13 | (correlation,nonlinearautocorr) | 77.38% |
| [187] | AC_nl_24 | (correlation,nonlinearautocorr) | 77.38% |
| [188] | AC_nl_135 | (correlation,nonlinearautocorr) | 77.38% |
| [192] | AC_nl_124 | (correlation,nonlinearautocorr) | 77.38% |
| [193] | AC_nl_134 | (correlation,nonlinearautocorr) | 77.38% |
| [196] | AC_nl_22 | (correlation,nonlinearautocorr) | 77.38% |
| [202] | AC_nl_022 | (correlation,nonlinearautocorr) | 77.38% |
| [208] | AC_nl_002 | (correlation,nonlinearautocorr) | 77.38% |

| | | | |
|---|---|---|---|
| [213] | AC_nl_012 | (correlation,nonlinearautocorr) | 77.38% |
| [214] | AC_nl_013 | (correlation,nonlinearautocorr) | 77.38% |
| [216] | AC_nl_014 | (correlation,nonlinearautocorr) | 77.38% |
| [221] | AC_nl_035 | (correlation,nonlinearautocorr) | 77.38% |
| [230] | AC_nl_122 | (correlation,nonlinearautocorr) | 77.38% |
| [531] | CO_glscf_1_1_3 | (correlation,glscf) | 77.38% |
| [536] | CO_glscf_1_2_2 | (correlation,glscf) | 77.38% |
| [543] | CO_glscf_1_5_3 | (correlation,glscf) | 77.38% |
| [553] | CO_glscf_2_2_1 | (correlation,glscf) | 77.38% |

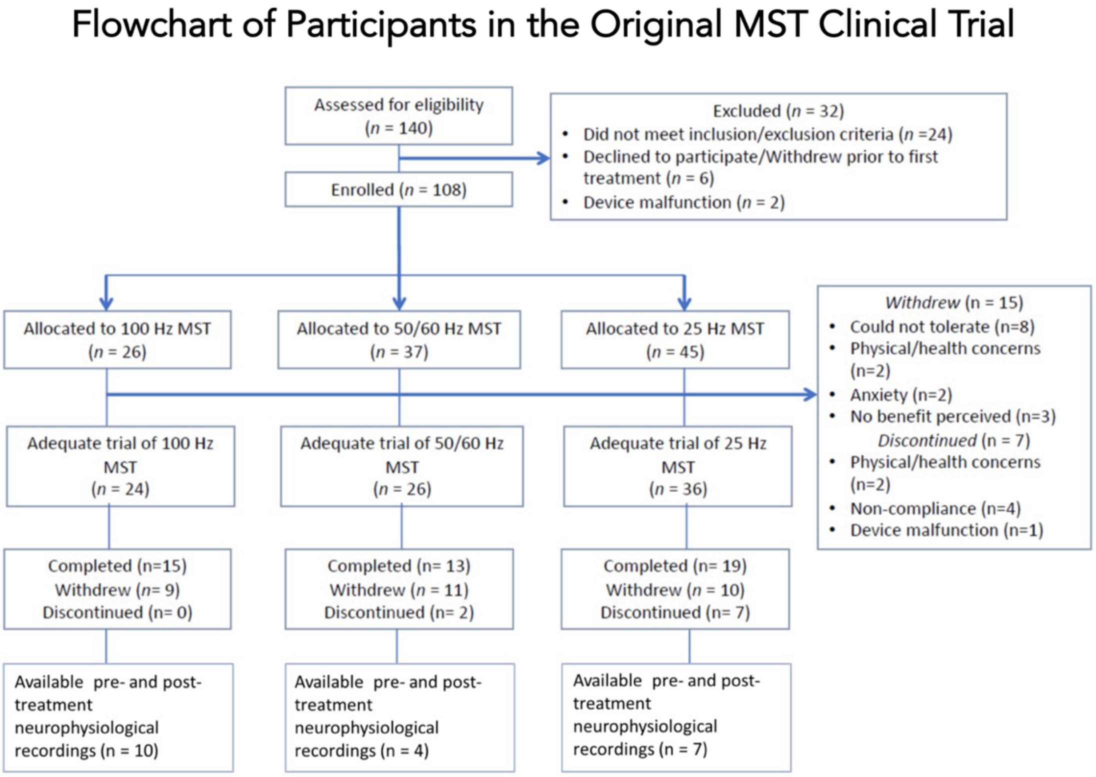


**Figure S1:** CONSORT diagram of the original MST clinical trial [described in detail in 5]. Of the patients who received an adequate course of treatment, 21 had complete, useable, neurophysiological data sets recorded at baseline and following the course of MST treatment. Note, no diagram is present for the ECT dataset, as this was from an observational study of the EEG correlates of stimulation efficacy in depression, and was not a registered clinical trial (see: [4]).

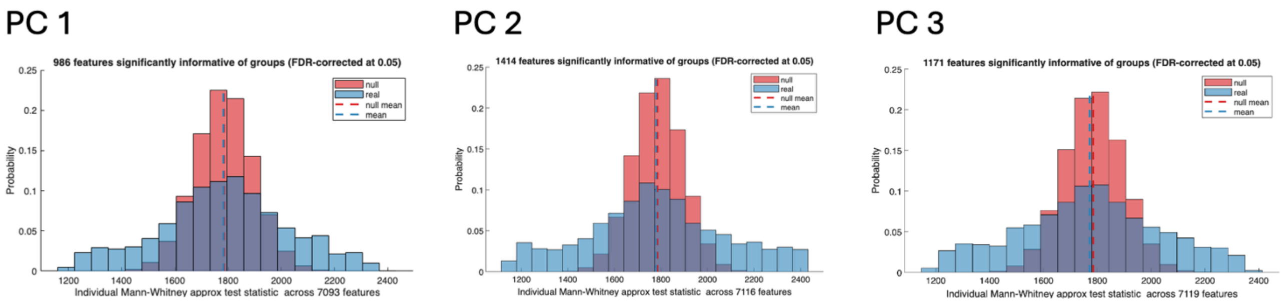


**Figure S2:** Distributions of feature-wise Mann-Whitney statistics for each EEG principal component (PC). For each PC, we compared pre- and post-treatment time-series features using the approximate Mann-Whitney U statistic implemented via *hctsa* (1,000 label permutations per feature). Red histograms show test statistics for the true group labels. Blue histograms show the pooled null distribution across all permutations. Vertical dashed lines indicate the mean of each distribution. For PC1, 986 of 7,093 features; for PC2, 1,414 of 7,116 features; and for PC3, 1,171 of 7,119 features exhibited statistics more extreme than the pooled null after false discovery rate correction at q=0.05.

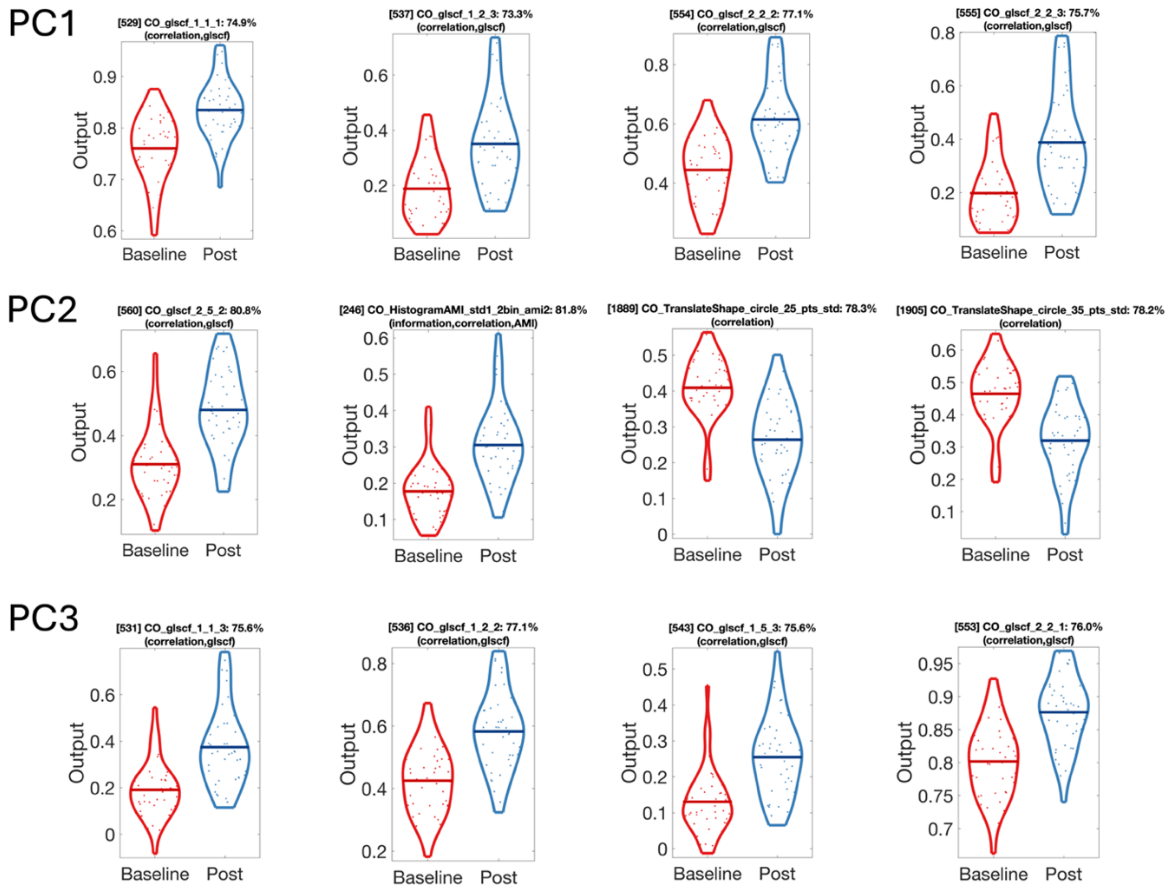


**Figure S3:** Violin plots at baseline and post-treatment showing four example features for each of the top three PCs capturing properties related to correlation within the time-series. Values were higher following treatment, broadly suggestive of greater temporal dependencies in the data. Two notable exceptions are features 1889 and 1905 for PC2, which relate to standard deviation within the 'CO_TranslateShape_circle' feature. This particular feature analyses the time-series by moving a geometric shape (in this case, a circle) along the data points and counting the number of points that fall within the circle at each position, thus providing insight into local density and the distribution of the data in the time series. The standard deviation reflects how much the counts vary as the circle moves across the time series. Lower variability, as noted post-treatment, might therefore reflect lower variability in the signal, possibly due to increased slow activity in the time series.

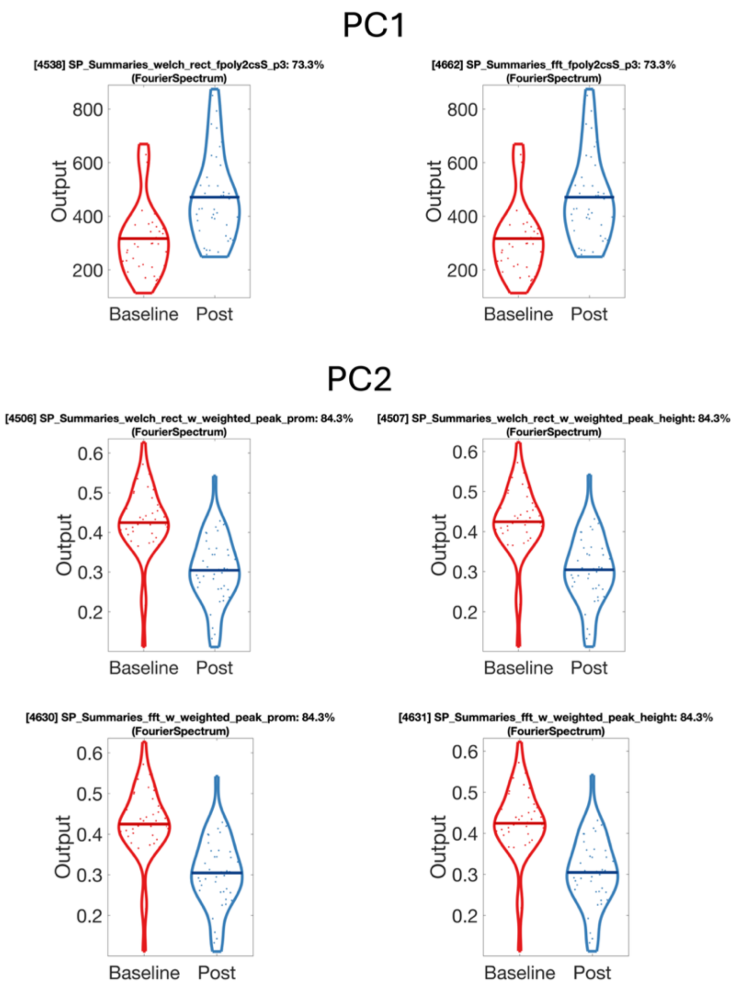


**Figure S4:** Violin plots of features related to the power spectra. For PC1, the features ‘SP_summaries_welch_rect_fpoly2csS_p3’ and ‘SP_Summaries_fft_fpoly2csS_p3’ had higher values following treatment compared to baseline. This indicates greater power in the EEG signal post-treatment is concentrated at lower frequencies (determined after transforming the signal into the frequency domain using either the Welch method [left], or FFT [right]). Similarly, there are lower values for both *peak prominence* and *peak height* (again, after either a Welch [middle row] or FFT [bottom row] transform). As can be seen, results are extremely similar between the Welch and FFT transformed data. Lower peak prominence suggests that peaks with higher prominence occur at lower frequencies on the power spectrum. Additionally, lower peak height indicates that higher amplitude peaks are occurring at lower frequencies following treatment. Together, these findings would be largely consistent with an EEG signal that has increased slow-wave activity.

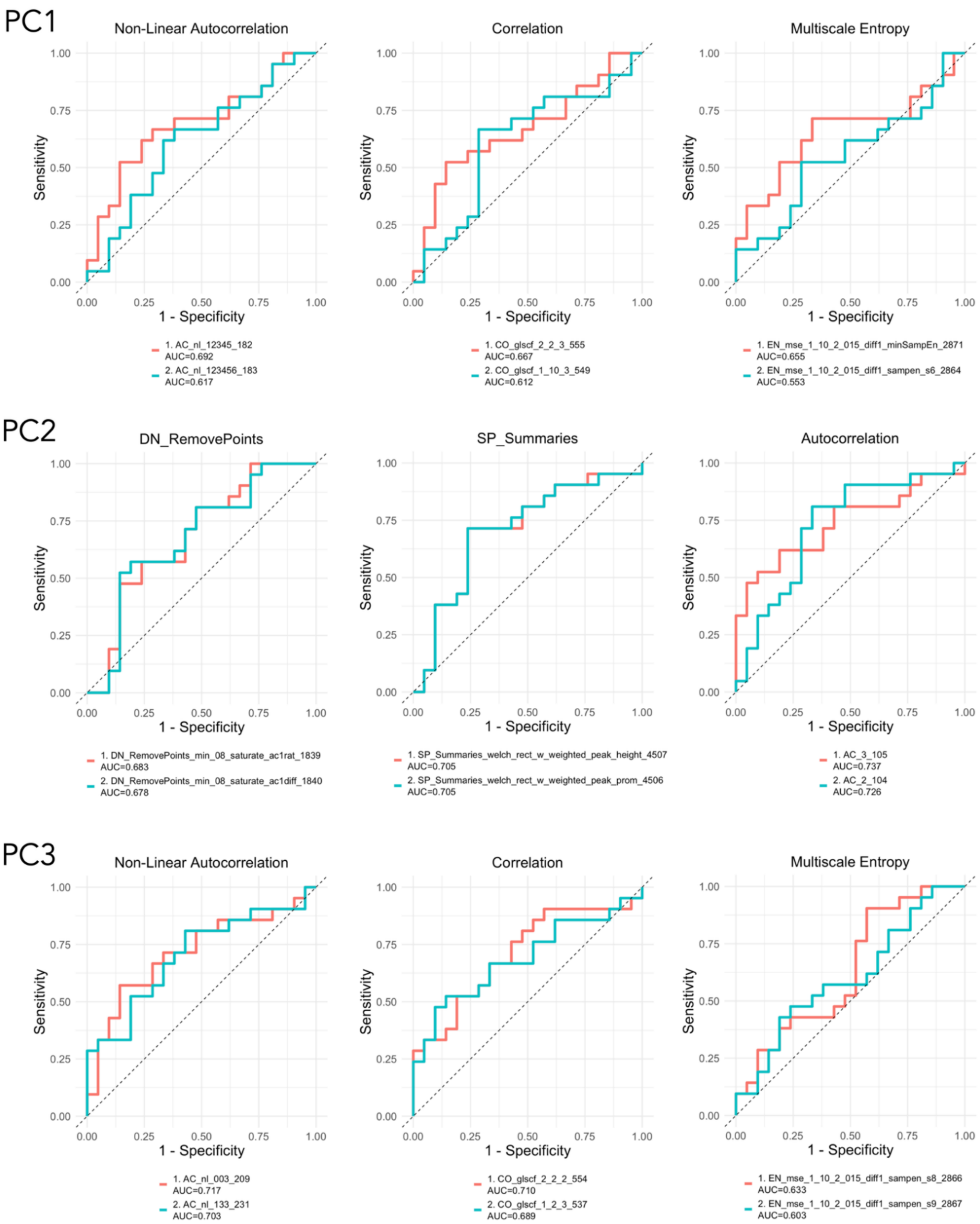


**Figure S5:** Receiver operating characteristic (ROC) curve plots showing the performance of top performing representative features at baseline (two features per curve) from each principal component (PC1-PC3) in predicting treatment responder status. Each panel illustrates features from different feature classes (e.g., non-linear autocorrelation, correlation, multiscale entropy), with curves plotting sensitivity (true positive rate) against 1 - specificity (false positive rate). Area under the curve (AUC) values for each feature are indicated in the legend within each panel.

**Table S2:** Results of the receiver operating characteristic analyses

| PC | Feature | AUC | CI (low) | CI (high) | Sensitivity | Specificity | p-value | $p_{FDR}$ |
|---|---|---|---|---|---|---|---|---|
| 1 | AC_nl_12345_182 | 0.69 | 0.53 | 0.86 | 0.67 | 0.71 | 0.034 | 0.072 |
| 1 | AC_nl_123456_183 | 0.62 | 0.44 | 0.79 | 0.67 | 0.62 | 0.195 | 0.234 |
| 1 | CO_glscf_2_2_3_555 | 0.67 | 0.50 | 0.84 | 0.52 | 0.86 | 0.213 | 0.240 |
| 1 | CO_glscf_1_10_3_549 | 0.61 | 0.43 | 0.79 | 0.67 | 0.71 | 0.064 | 0.096 |
| 1 | EN_mse_1_10_2_015_diff1_minSampEn_2871 | 0.66 | 0.48 | 0.83 | 0.71 | 0.67 | 0.554 | 0.554 |
| 1 | EN_mse_1_10_2_015_diff1_sampen_s6_2864 | 0.55 | 0.37 | 0.73 | 0.52 | 0.71 | 0.085 | 0.118 |
| 2 | AC_3_105 | 0.74 | 0.58 | 0.89 | 0.62 | 0.81 | 0.009 | 0.062 |
| 2 | AC_2_104 | 0.73 | 0.57 | 0.89 | 0.81 | 0.67 | 0.012 | 0.062 |
| 2 | DN_RemovePoints_min_08_saturate_ac1rat_1839 | 0.68 | 0.52 | 0.85 | 0.81 | 0.52 | 0.048 | 0.079 |
| 2 | DN_RemovePoints_min_08_saturate_ac1diff_1840 | 0.68 | 0.51 | 0.85 | 0.57 | 0.81 | 0.043 | 0.077 |
| 2 | SP_Summaries_welch_rect_w_weighted_peak_height_4507 | 0.71 | 0.54 | 0.87 | 0.71 | 0.76 | 0.023 | 0.062 |
| 2 | SP_Summaries_welch_rect_w_weighted_peak_prom_4506 | 0.71 | 0.54 | 0.87 | 0.71 | 0.76 | 0.023 | 0.062 |
| 3 | AC_nl_003_209 | 0.72 | 0.56 | 0.88 | 0.57 | 0.86 | 0.016 | 0.062 |
| 3 | AC_nl_133_231 | 0.70 | 0.54 | 0.86 | 0.81 | 0.57 | 0.024 | 0.062 |
| 3 | CO_glscf_2_2_2_554 | 0.71 | 0.55 | 0.87 | 0.81 | 0.52 | 0.036 | 0.072 |
| 3 | CO_glscf_1_2_3_537 | 0.69 | 0.52 | 0.86 | 0.52 | 0.86 | 0.020 | 0.062 |
| 3 | EN_mse_1_10_2_015_diff1_sampen_s8_2866 | 0.63 | 0.46 | 0.81 | 0.90 | 0.43 | 0.252 | 0.267 |
| 3 | EN_mse_1_10_2_015_diff1_sampen_s9_2867 | 0.60 | 0.43 | 0.78 | 0.48 | 0.76 | 0.141 | 0.181 |

*Note:* AUC = area under the curve; CI = confidence interval; PC = principal component.

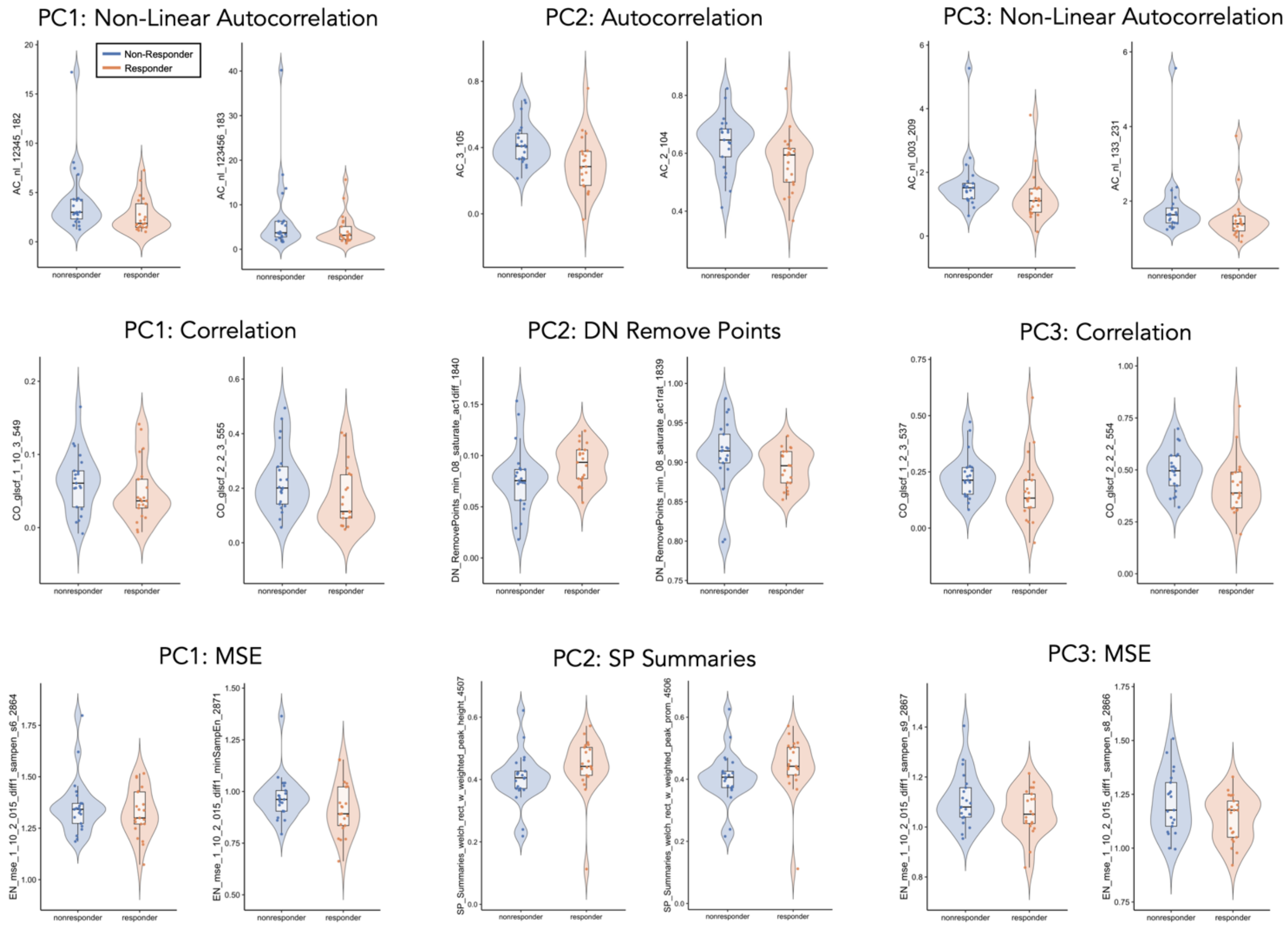


**Figure S6:** Plots of representative top-performing features from each feature cluster from each of the three principal components that were assessed using ROC curves (PCs; total of 18 features across the three PCs). Plots show values at baseline for responders and non-responders to treatment.
